# Machine Learning for Longitudinal Brain-Age Prediction

**DOI:** 10.64898/2025.12.10.25341964

**Authors:** Manuel Wegmann, Melanie Ganz, Jonas E Svensson, Pontus Plavén-Sigray, Ruben P. Dörfel

**Affiliations:** Neurobiology Research Unit, Rigshospitalet, Copenhagen, DK; Department of Computer Science, University of Copenhagen, Copenhagen, DK; Centre for Psychiatry Research, Department of Clinical Neuroscience, Karolinska Institutet & Stockholm Health Care Services, Region Stockholm, SE

## Abstract

Cross-sectional brain age models have demonstrated high accuracy and reliability for predicting chronological age based on structural brain features derived from single MRI scans. However, these models cannot separate baseline variation from true aging-related changes or noise. Longitudinal models address this limitation by predicting inter-scan intervals from paired MRI scans, controlling for baseline factors through repeated measurements. Using OASIS-3 data, we compare a cross-sectional 3D CNN against three longitudinal architectures for predicting inter-scan intervals: LILAC (Siamese neural network), LILAC+ (enhanced Siamese network with multi-layer perceptron), and AM (variational autoencoder). Longitudinal models substantially outperformed the cross-sectional approach, with LILAC+ achieving best performance (MSE = 1.97 years^2^, MAE = 0.99 years, r = 0.86, R^2^ = 0.71). Our results suggest that direct modeling of longitudinal change is more effective at capturing individual aging trajectories than deriving intervals from cross-sectional predictions.

## 1. INTRODUCTION

Aging, the accumulation of molecular and cellular damage over time, predisposes individuals to neurodegeneration, cognitive decline, and dementia [1]. To assess individual disease risk and develop effective interventions, reliable biomarkers of aging are necessary.

*Brain age*, a proposed biomarker inferred from structural MRI scans of the brain, estimates the apparent age using machine learning models trained on healthy individuals [2]. These models assume biological age (BA) aligns with chronological age (CA) in healthy brains and thus characterize a normative aging pattern. In order for a brain age biomarker to be clinically useful, it should predict future neurodegeneration and cognitive decline before symptoms manifest.

Although brain age models have shown high accuracy and reliability [3], brain age does not predict disease conversion, gray matter atrophy, or cognitive decline [4], likely because cross-sectional data cannot distinguish between confounding baseline effects, aging-related biology, and noise [5].

To address these limitations, recent approaches have suggested using multiple MRI scans from the same participants [6, 7]. Instead of predicting age at specific timepoints, these methods predict inter-scan intervals, learning aging-related changes while controlling for congenital baseline factors. In this study, we extend the proposed framework [7] and compare cross-sectional models against two longitudinal encoding architectures: a Siamese neural network and a pretrained variational autoencoder.

## 2. METHODS

### 2.1 Problem Formulation

A longitudinal dataset *D* in this study consists of *N* subjects, each of whom has completed a baseline MRI scan and a follow-up scan. Subject *i* underwent their baseline and follow-up scans at time points 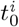 and 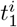, where corresponding MRIs are denoted by 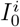 and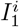.The task is to estimate the inter-scan time interval, denoted by:

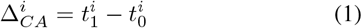

For healthy subjects, it is assumed that the change in brain age between *t*_0_ and *t*_1_, denoted by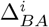, closely aligns with the change in chronological age 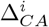, i.e., the time passed or inter-scan interval. Models are trained to learn a function *f* such that:

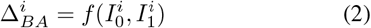

The model is optimized by minimizing a loss functional over the dataset *D* of healthy subjects:

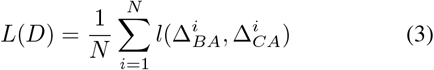

Here, *l* refers to the point-wise loss function, which in this study is defined as the squared error.

### 2.2 Data

We used 3T brain MRI data from the OASIS-3 study (Open Access Series of Imaging Studies) [8]. Baseline demographics are provided in Table 1. For training, we included only cognitively normal individuals with at least two scans. For participants with more than two scans, all possible scan pairs with positive time intervals were generated, resulting in 2039 scan pairs. The inter-scan interval ranges from 0 to 12.3 years (mean: 4.18 years).

**Table 1.** Baseline demographic characteristics of cognitively normal participants from the OASIS-3 cohort. Scans denotes the average number of MRI sessions completed per participant, and Dur. represents the mean duration between the first and last MRI session. Participants with only one session have a duration of 0 years.

| N | Sex (M/F/U) | Age [years] | Scans | Dur. [years] |
| --- | --- | --- | --- | --- |
| 1000 | 327/461/212 | 68.7 ± 9.8 | 2.35 | 3.42 |

MRI images were preprocessed and segmented using FastSurfer (cuda-v2.4.2) [9]. Brain-masked and bias-field-corrected images were registered to the template space (MNI152NLin2009cAsym) using an affine transformation.

### 2.3. Models

Figure 1 provides a conceptual overview of a model architecture specifically designed for longitudinal MRI data: a pair of scans is passed through a shared encoder to extract feature representations, which are then subtracted to form a difference vector in feature space and passed through a prediction layer to estimate the target inter-scan interval. The following models were implemented for this study:

**Fig. 1.**
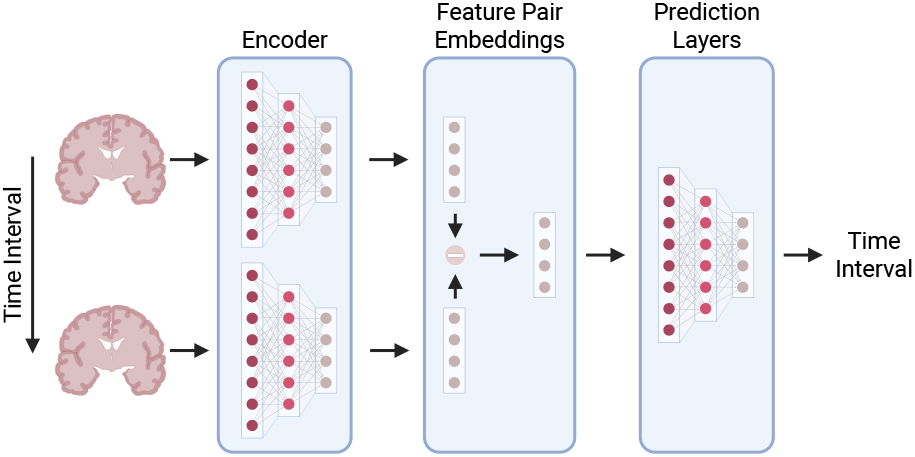
Schematic overview of model architecture for longitudinal brain age task.

#### Baseline reference

A linear regression model predicting inter-scan intervals from baseline age serves as the comparison standard. Models not outperforming this baseline will have failed to learn meaningful imaging features.

#### CS CNN

This cross-sectional model uses a single-scan CNN encoder with identical architecture to LILAC. Features are concatenated with sex and passed through a linear layer to predict chronological age. The model was trained to predict chronological age using 1,756 MRI scans from cognitively normal participants in the OASIS-3 dataset via 5-fold crossvalidation. After training, longitudinal estimates are obtained on the validation set by subtracting the baseline age estimate from the follow-up age estimate.

#### LILAC [7]

A Siamese architecture processes paired scans through a shared CNN encoder. Feature differences are concatenated with sex and passed through a linear layer to predict inter-scan intervals.

#### LILAC+

Extends LILAC by replacing the linear prediction layer with a two-layer multilayer perceptron (2,048 and 512 neurons with ReLU activation), followed by a linear layer with softplus activation to capture non-linear relationships.

#### AM

An autoencoder-based model using MedVAE [10], a variational autoencoder for medical imaging that was pretrained on multiple datasets, including OASIS-3. Latent representations (downsampled 4× spatially) from paired scans are subtracted, concatenated with sex, and passed through linear regression to predict inter-scan intervals.

### 2.4 Training

All models were implemented in PyTorch [11] and trained on a single NVIDIA V100 GPU. The code is publicly available at https://github.com/manuelwegmann/longitudinal_brainage.

Models were trained using MSE loss and the Adam optimizer [12] with a learning rate decay. Hyperparameters were determined through preliminary test runs.

We used stratified 5-fold cross-validation based on baseline age to ensure comparable age distributions across folds. Scan pairs were generated only after fold assignment. Predictions from all folds were concatenated to evaluate overall performance on the entire dataset.

### 2.5 Evaluation

Model performance was evaluated using four metrics comparing predicted and true inter-scan intervals. Mean Squared Error (MSE) and Mean Absolute Error (MAE) quantify prediction accuracy, with MSE penalizing larger errors more heavily. Pearson’s r measures the linear correlation between predicted and observed intervals. The coefficient of determination (R^2^) indicates the proportion of variance in true intervals explained by the model. While related, Pearson’s r reflects the strength of linear dependence independent of bias, and R^2^ measures overall goodness of fit.

## 3. RESULTS

Table 2 summarizes the main results, showing performance metrics across all evaluated models. The baseline linear regression model had an MSE of 6.61 years^2^, MAE of 2.07 years, and *R*^2^ values around zero, indicating that it failed to explain the variance in the target variable. It showed a statistically significant but weak correlation between predictions and targets, with a Pearson’s *r* value of 0.18.

**Table 2.**
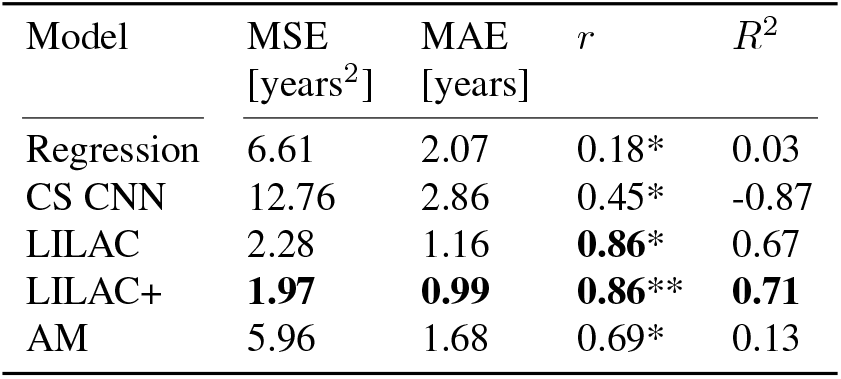
Summary statistics of the various models. Pearson’s r between the predicted and actual inter-scan interval is denoted by *r*, where * indicates an associated p-value of *<* 10^−6^. *R*^2^ denotes the coefficient of determination.

The cross-sectional CNN model (CS CNN) showed an improvement in correlation over the regression model, achieving a statistically significant, moderate correlation of *r* = 0.45. However, its overall accuracy was worse than the baseline model with an MSE of 12.76 years^2^ and an MAE of 2.86 years, indicating high variability in prediction accuracy. Similarly, the *R*^2^ value of −0.87 suggests that this crosssectional approach fails to capture the variability in the target values.

LILAC achieved improved performance across all metrics, with an MSE of 2.28 years^2^ and an MAE of 1.16 years. It also achieved a statistically significant, high Pearson’s r of *r* = 0.86 and a *R*^2^ value of 0.67, indicating that the model explains a moderate proportion of the variance in the target variable.

Building upon LILAC, its more complex variant LILAC+ was the most accurate model overall, achieving an MSE of 1.97 years^2^ and MAE of 0.99 years. The correlation remained high, with *r* values of 0.86. The *R*^2^ score improved slightly to 0.71, indicating that this enhanced model explains a slightly larger proportion of variance in the target variable compared to the original LILAC.

The autoencoder-based model (AM) performed worse than the LILAC variants, with an MSE of 5.96 years^2^ and MAE of 1.68 years. Similarly, Pearson’s *r* was lower at 0.69, and the *R*^2^ score was 0.13, indicating only moderate correlation and a small proportion of variance explained. Nonetheless, this represents a noticeable improvement over the cross-sectional model and baseline regression model.

The scatter plots in Figure 2 shows that the LILAC and LILAC+ models exhibit the strongest linear fit between the predictions and targets and relatively low variability. AM also captures the underlying trend, though the predictions display greater variability. In contrast, CS CNN appears unable to capture the data distribution effectively and tends to severely underestimate larger target values.

**Fig. 2.**
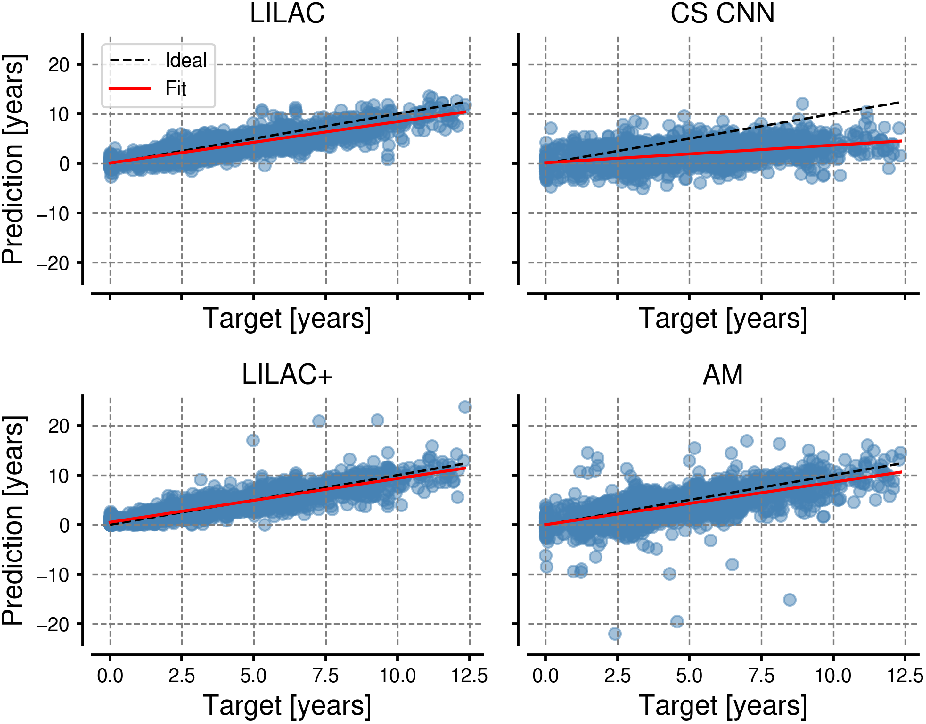
Scatter plots of predicted versus true time intervals between scans for all models. For each model, a regression line is fitted alongside the ideal diagonal line representing perfect agreement between predictions and targets.

## 4. DISCUSSION

In this study, we evaluated and compared cross-sectional and longitudinal MRI-based models for predicting inter-scan brain age intervals in healthy adults, examining how different architectural choices impact the accuracy of capturing aging-related changes.

The longitudinal models were able to predict inter-scan time accurately, though performance varied across architectures. Notably, all longitudinal models outperformed the baseline linear regression model across all metrics, suggesting that the longitudinal models learned relevant image-based features related to aging. Particularly, LILAC and LILAC+ demonstrated high accuracy, indicating their capability to accurately track inter-scan time intervals from brain MRIs in healthy adults. These findings are consistent with previous results [7], where LILAC outperformed naive baseline models for the longitudinal brain age task, achieving an MSE of 1.83, an MAE of 1.09, and a Pearson’s *r* of 0.85. In contrast to the longitudinal models, the cross-sectional model failed to accurately track aging in healthy adults, performing worse than baseline linear regression on both MAE and MSE. Further, its *R*^2^ value of −0.87 indicates that the model fails to explain the variance in the target.

Notably, CS CNN shares much of LILAC’s architecture but predicts chronological age from single scans and derives intervals by taking the difference. Given these similarities, the observed performance gap suggests substantial methodological advantages to directly modeling longitudinal change, indicating that such an approach may be more effective for accurately capturing brain aging trajectories under otherwise comparable conditions. It should be emphasized that CS CNN was intentionally designed to be comparable to LILAC, potentially placing it at a disadvantage. Therefore, future studies should include state-of-the-art cross-sectional models to evaluate performance gaps between longitudinal and cross-sectional architectures. Nonetheless, the findings in this study strongly support the conclusion that, within a comparable framework, longitudinal modeling approaches are more effective at capturing the aging process in healthy adults.

Despite these promising results for longitudinal models, the *R*^2^ values (AM: 0.19, LILAC: 0.67, LILAC+: 0.71) show that, while a considerable portion of variance in the data was explained by LILAC and LILAC+, there is still substantial unexplained variability, especially for AM. This implies the presence of underlying patterns that have not yet been captured and which need to be further addressed. Possible approaches include constructing a larger dataset to extract more distinct aging-related features or incorporating additional demographic information and clinical variables, such as multimodal imaging data.

This study focused on evaluating the accuracy of different model architectures in tracking age progression in cognitively normal adults. To validate their utility as biomarkers for detecting accelerated aging and disease risk, future work should examine model performance across cohorts with varying degrees of neurodegeneration and explore associations between model predictions and clinical markers, such as cognitive test scores.

The performance difference between LILAC and AM suggests that simpler, task-specific feature extraction may be more effective than general-purpose autoencoders like MedVAE. While AM’s autoencoder structure enables interpretability through image reconstruction, competitive performance likely requires task-specific adaptation such as fine-tuning and combined loss functions [13].

In summary, longitudinal brain age models, particularly LILAC and LILAC+, reliably predict inter-scan intervals and outperform a comparable cross-sectional approach, highlighting their potential as biomarkers of brain health. Future work should evaluate these models on larger, multi-site datasets, assess their ability to predict clinical outcomes across diverse populations, and ensure within-subject stability of predicted trajectories.

## 5. COMPLIANCE WITH ETHICAL STANDARDS

This research study was conducted retrospectively using human subject data made available in open access by OASIS-3 [8]. Ethical approval was not required as confirmed by the license attached with the open access data.

## Data Availability

All data produced in the present study are available upon reasonable request to the authors

https://github.com/manuelwegmann/longitudinal_brainage

## 6. ACKNOWLEDGMENTS

No funding was received for this study, and the authors have no conflicts of interest to disclose. Computations were enabled by resources from the National Academic Infrastructure for Supercomputing in Sweden (NAISS), partially funded by the Swedish Research Council (grant no. 2022-06725).

